# Decreased central corpus callosum volume is associated with repetitive behaviours and motor difficulties in autistic children

**DOI:** 10.1101/2024.06.20.24309217

**Authors:** Gaia Scaccabarozzi, Denis Peruzzo, Filippo Arrigoni, Silvia Busti Ceccarelli, Laura Villa, Elisa Mani, Eleonora Maggioni, Paolo Brambilla, Maria Nobile, Massimo Molteni, Alessandro Crippa

## Abstract

Along with the core characteristics of the condition, autistic individuals commonly experience motor coordination difficulties, potentially related to a reduced cortical connectivity. Being the largest human commissure, the corpus callosum (CC) plays an essential role in interhemispheric connectivity and has been often involved among autistic atypicalities. This study aimed to investigate the volumes of corpus callosum subregions in a group of drug-naïve, autistic children and to explore its possible associations with both core features and motor coordination skills. Thirty-five autistic children (2.5-12 years) were compared with a group of 35 closely IQ-matched, non-autistic peers. CC was identified and segmented into five subregions using Freesurfer. Callosal volumes were compared between the two groups and correlated with parental ratings of core autistic features as assessed by the Social Responsiveness Scale and with motor features as assessed by the Developmental Coordination Disorder Questionnaire. Associations between CC volume and Autism Diagnostic Observation Schedule scores were also explored in autistic participants. Autistic children showed a reduced volume of the central segment of the CC, in the context of a comparable CC total volume. This reduction appeared to be correlated with symptoms of restricted and repetitive behaviours in autistic children, and to parental ratings of autistic mannerisms and motor skills across participants. These findings expand the current knowledge about the neural mechanisms underlying autism, suggesting that the reduced connectivity through the CC might have implications for both core and motor features of autistic individuals.

**Lay Summary:** Differences in brain development have been widely outlined in autism. Exploring brain scans of 35 autistic and non-autistic children aged 2.5-12 years and closely matched for cognitive functioning, we found that the central part of the corpus callosum was smaller for the autistic group. This reduction was associated with the level of restricted and repetitive behaviours in autistic children, and to parental ratings of autistic mannerisms and motor coordination skills across participants. This work offers new empirical evidence that interhemispheric connectivity is atypical in autism and that the corpus callosum can be involved in the manifestation of both core and motor characteristics of autistic children.

## 1. INTRODUCTION

Autism is a complex neurodevelopmental condition mainly characterised by atypical social communication together with focused and circumscribed behaviours and interests (Lord et al., 2020; Wright, Spikins, & Pearson, 2020). Along with these core characteristics, autistic individuals often report motor coordination difficulties on daily living skills (Gowen, Earley, Waheed, & Poliakoff, 2023) and motor impairments/delays have been extensively described in a meaningful subpopulation of autistic children (∼87%; Bhat, 2020). Some recent studies (e.g., Lidstone & Mostofsky, 2021, Wang et al. 2022) proposed that the atypical sensorimotor manifestations described in autism could stem from a less effective ability to integrate multiple information to guide ongoing (motor) behaviours. Indeed, autism-related differences are particularly apparent on tasks such as ball catching and motor imitation, which require efficient sensory-motor integration (De Francesco, Morello et al., 2023; Lainé, Rauzy, Tardif, & Gepner, 2011; Whyatt & Craig, 2012), and whole-body coordination tasks, where the integration of motor commands from the two hemispheres is needed (Chen et al., 2019). Furthermore, previous research has leveraged the sensorimotor domain to decompose the autism heterogeneity (Craig et al., 2021; Mandelli et al., preprint) and support its identification (De Francesco, Morello et al., 2023; Lidstone, Mostofsky, & Ewen, preprint), but also to suggest potential alterations in underlying brain systems (Lepping et al., 2022; Lidstone, Rochowiak, Mostofsky, & Nebel, 2021; Oldehinkel et al., 2019).

In this respect, differences in brain development have been widely outlined in autism. Increased total brain volume is a well-replicated finding, especially in early childhood (Sacco, Gabriele, & Persico, 2015), as well as alterations in grey and white matter volume of multiple discrete brain regions (Kangarani-Farahani, Izadi-Najafabadi, & Zwicker, 2022; Pagnozzi, Conti, Calderoni, Fripp, & Rose, 2018). Given the widespread spatial distribution of the neural atypicalities, autism research has also targeted large-scale patterns of connectivity between different neural networks (Geschwind & Levitt, 2007; Vasa, Mostofsky, & Ewen, 2016), with evidence of coexisting over- and under-connectivity in distinct topographies and/or developmental stages (Di Martino et al., 2014). Moreover, peculiarities in connectivity have been linked to both core and non-core features of autism (Just, Keller, Malave, Kana, & Varma, 2012), including visuo-motor coordination (Lepping et al., 2022).

The corpus callosum (CC) includes most of the fibres connecting right and left hemispheres (Bellani, Calderoni, Muratori, & Brambilla, 2013), thus mediating the communication across contralateral brain regions (i.e., interhemispheric connectivity). On the basis of decades of imaging data (see Bellani, Calderoni, Muratori, & Brambilla, 2013; Valenti et al., 2020) and insights coming from patients with callosal agenesis (Booth, Wallace, & Happé, 2011; Romaniello et al., 2017), it has been suggested that the CC could be implicated in autism. However, studies in the literature offered contrasting findings about CC volume in autism. Two meta-analyses reported indeed a reduction of CC volume in autistic individuals, with an effect size of 0.5 (Frazier & Hardan, 2009; Lefebvre, Beggiato, Bourgeron, & Toro, 2015). Conversely, a number of studies examining the Autism Brain Imaging Data Exchange (ABIDE) database (∼1000 subjects) provided mixed evidence. Lefebvre and colleagues did not find any difference between autism and non-autism groups in either total or segmented CC volumes (Lefebvre, Beggiato, Bourgeron, & Toro, 2015), whereas Haar and others and Loomba and colleagues found a reduction limited to the central CC volume in autism (Haar, Berman, Behrmann, & Dinstein, 2016-; Loomba, Beckerson, Ammons, Maximo, & Kana, 2021). Nevertheless, Li and others reported decreased volume of CC anterior and posterior, but not central (Li et al., 2019). While these studies considered different subsets of participants taken from the ABIDE database, there might be other reasons to these discrepant findings, as multiple factors, such as age, magnetic resonance imaging (MRI) magnet strength, MRI scanning site, sex, and intelligence quotient (IQ) can influence, not necessarily in a linear way, CC volumes’ values (Frazier & Hardan, 2009; Lefebvre, Beggiato, Bourgeron, & Toro, 2015). With specific respect to the effect of age, a lab specific whole-brain MRI study including 1769 subjects aged 0–32 years found that several CC’s subregions showed the largest group wise differences between autistic and non-autistic participants, specifically in the age range 15-20 years (Levman et al., 2018). Interestingly, the volume reduction of these CC’s regions was apparent from 5 years of age, while younger autistic toddlers presented larger CC volumes (in the context of increased total brain volume) when compared to non-autistic controls (Levman et al., 2018).

Atypicalities in CC volumes have been also linked to autism core features, with previous research showing positive associations between larger callosal size and lower levels of autistic features as assessed by the Autism Diagnostic Observation Schedule (Prigge et al., 2021; Giuliano et al., 2018) or as rated by parents (Hardan et al., 2009; Loomba, Beckerson, Ammons, Maximo, & Kana, 2021; Wolff et al., 2015). CC volumes have been also associated with non-core features of autism, such as higher scores on standardised measures of IQ (Prigge et al., 2013) or better executive functioning (Keary et al., 2009). Despite the crucial role of the CC for both motor coordination and sensory integration (Moes, Schilmoeller, & Schilmoeller, 2009; Rademaker et al., 2004), initial investigations in autism did not find significant association between atypical CC volumes and direct assessments of motor skills (assessed with the Zurich Neuromotor Assessment in Freitag et al., 2009 and with the Movement Assessment Battery for Children 2 in Hanaie et al., 2014).

The present study sought to extend the results of these two latter studies in a sample of drug-naïve autistic participants, presenting lower scores on standardised measures of IQ and younger ages. We first investigated whether a group of well-characterised autistic children would demonstrate a reduced CC volume in comparison with a group of closely IQ-matched, non-autistic peers. We further aimed to examine how CC size is associated with both core features of autism and caregiver reports on motor skills. Since the central CC is the largest human commissural section connecting homologous cortical regions involved in motor planning and execution (Hofer & Frahm, 2006; Russo et al., 2022), we hypothesised that the volume of the central CC would be related to the motor characteristics of our participants.

## 2. METHODS

### 2.1 Participants

Seventy children aged 2.5-12 years participated in this study, including 35 autistic children and 35 IQ-matched, non-autistic peers. Participants in the autism group were recruited from in and outpatient populations of the Scientific Institute, IRCCS Eugenio Medea (Bosisio Parini, Italy), a tertiary referral clinical centre. At admittance, autistic children were diagnosed according to Diagnostic and Statistical Manual of Mental Disorders (5th ed.; DSM-5) criteria by a multidisciplinary team including a medical doctor specialised in child neuropsychiatry with experience in autism, an experienced child psychologist, and a speech therapist. The diagnoses were supported by using the Autism Diagnostic Observation Schedule–Second Edition (ADOS-2; Lord et al., 2012). Intellectual disability (IQ<70) was reported in 17 autistic participants using an appropriate developmental/intelligence scale. Furthermore, based on the clinical assessment, children in the autism group received the following additional diagnoses according to the DSM-5 criteria: attention-deficit hyperactivity disorder (N=2), specific language disorder (N=2), specific learning disorder (N=2), and developmental coordination disorder (N=1). The participants in the comparison group were selected to be closely matched to the autistic children on an individual basis for intelligence quotient (IQ). We decided to match children by IQ based on previous evidence in autism of a significant association between cognitive functioning and CC volumes. Twenty-seven of the children in the comparison group had been initially referred to our Institute for suspected developmental delays. After having undergone a comprehensive diagnostic evaluation (see above), autism was thoroughly ruled out for all the cases. Fifteen of the 27 referred children received a diagnosis of idiopathic intellectual disability after a negative assessment for underlying causes. Other co-occurring diagnoses for these participants were childhood obesity (N=1), specific language disorder (N=1), specific learning disorder (N=1). The remaining 8 participants in the comparison group had been recruited as part of various research studies at our Institute from the general population by local paediatricians or schools near the institute. They had no previous history of social/communicative disorder, other developmental concerns, or family history of any neurodevelopmental condition. Exclusion criteria for all participants were using any stimulant or non-stimulant medication affecting the central nervous system, having a well-defined genetic disorder, having a significant sensory impairment (e.g., blindness, deafness), presence of contraindications to MRI exam or having abnormalities detected by MRI, suffering from chronic or acute medical illness. All participants were drug-naïve.

The present research complies with the ethical standards of the 1964 Declaration of Helsinki and later amendments and was approved by the Ethics Committee of our Institute “Comitato Etico IRCCS E. Medea—Sezione Scientifica Associazione La Nostra Famiglia” (Prot. N.33/18—CE). All of the participants’ parents or legal guardians gave informed written consent before participation.

### 2.2 Measures

The IQ level was assessed with different scales, according to the participants’ developmental level: Griffiths Mental Development Scales (Griffiths, 1970), Wechsler Preschool and Primary Scale of Intelligence (WPPSI-III; (Wechsler, 2002)), and Wechsler Intelligence Scale for Children–IV (Wechsler, 2012).

The Social Responsiveness Scale (SRS; Constantino & Gruber, 2005) was used to assess the degree of autistic traits/symptoms as reported by parents. Its 65 items address reciprocal social behaviours and are grouped into five subscales: social awareness, social cognition, social communication, social motivation, and autistic mannerisms. For each item, answers are given on a 5-point Likert scale and higher scores indicate increased autistic traits. The five SRS subscale scores were considered as dependent measures.

The participants’ motor skills were measured using the parent-report Developmental Coordination Disorder Questionnaire (DCDQ; Wilson, Kaplan, Crawford, & Roberts, 2007), which provides an estimate of gross and fine motor functioning in children. The questionnaire is composed of 15 items addressing different subdomains of motor abilities, such as ball skills, complex motor coordination, and fine and general motor skills. For each item, parents rate the children’s degree of motor ability on a 5-point scale comparing it with peers of the same age. The three subscales of the DCDQ, addressing motor control, fine motor, and general coordination, respectively, contribute to determine a total score, with higher scores meaning better motor functioning. The three DCDQ subscale scores were recorded as dependent measures.

In the autistic group, raw social affect (SA) and restricted repetitive behaviours (RRB) domains scores and total raw scores from the ADOS (Lord et al., 2012) were separately converted into calibrated severity scores following the procedure by Gotham et al. (2009) and Hus et al. (2014). Indeed, calibrated severity scores provide comparability across individuals with different developmental levels (Gotham, Pickles, & Lord, 2009; Hus, Gotham, & Lord, 2014).

Lastly, familial socioeconomic status (SES) was coded according to the Hollingshead scale for parental employment (Hollingshead, 1975).

### 2.3 MRI data acquisition and processing

MRI data were acquired on a 3T Philips Achieva d-Stream scanner (Best, The Netherlands) with a 32-channel head coil. T1-weighted (T1W) data were acquired with a 3D Turbo Field Echo sequence with Field of View (FOV) = 256x256x175 mm^3^, voxel size = 1x1x1 mm^3^, Time of Repetition (TR) = shortest (∼8.1 ms), Time of Echo (TE) = shortest (∼3.7 ms) and Flip Angle (FA) = 8°. Image processing was performed using the Freesurfer image analysis suite (V6) (http://surfer.nmr.mgh.harvard.edu/) using the standard “recon-all” pipeline. Briefly, images were masked to remove non-brain tissue, registered to the Talairach space and segmented to extract the cortical grey matter, the white matter, and the deep grey matter structures (Fischl et al., 2004). After visual inspection and manual correction of the previous steps, surfaces of the white/grey and grey/cerebrospinal fluid borders were computed (Fischl & Dale, 2000) and registered to a spherical atlas (Fischl, Sereno, Tootell, & Dale, 1999) allowing a subject specific parcellation of the cerebral cortex according to the gyral and sulcal structure using a standard anatomical nomenclature (Destrieux, Fischl, Dale, & Halgren, 2010). Through the Freesurfer processing steps, CC was thus automatically identified and segmented into five equally spaced subregions for each subject: Anterior CC, Mid Anterior CC, Central CC, Mid Posterior CC and Posterior CC. Total CC volume was calculated as the sum of these five segment volumes for each participant. Total intracranial volume (TIV) was also estimated using Freesurfer including everything except the cerebellum and the brain stem.

### 2.4 Statistical analysis

Statistical analyses were performed using SPSS Statistics Software (Version 21). After checking for linear model assumptions, between-group differences for demographic variables (age, IQ, and SES) were analysed under a univariate analysis of variance (ANOVA), while a chi-square was performed to examine differences in sex distribution for the two groups. An ANOVA was then used to explore between-group differences in TIV. Between-group differences in CC were first tested with ANOVA. The statistical analysis of CC volumes (both total volume and the volumes of the 5 CC subregions) was then repeated using the analysis of covariance to control for possible confounding factors such as TIV and age (Prigge et al., 2021; Wang et al., 2024). Results were adjusted for multiple comparisons using a Bonferroni correction based upon the 6 statistical tests performed on the CC measures. Pearson’s correlations were conducted to examine the relationship between CC volumes and social communication/motor measures. Partial linear correlations were also conducted to explore the significance of these relationships while controlling for TIV and age. Lastly, Spearman’s correlation was performed to explore possible relationships between CC volumes and ADOS in the autistic group. Given the exploratory nature of these correlations, no correction was applied for multiple comparisons.

## 3. RESULTS

### 3.1 Sample description

Group characteristics are summarised in Table 1. The autistic and non-autistic groups were matched for IQ and balanced on SES and TIV; the groups also did not significantly differ in sex and age (all p > 0.1).

**Table 1.** Sample description. IQ= intelligence quotient; SES= familial socioeconomic status; TIV= total intracranial volume; ADOS= autism diagnostic observation schedule; SA= Social Affect; RRB= Restricted Repetitive Behaviours. a Chi-square; b ANOVA.

|  | **Autistic** | **Non-autistic** | **p-value** |
| --- | --- | --- | --- |
| **Sex (M:F)** | 30:5 | 25:10 | 0.145<sup>a</sup> |
| **Age (years)**<br>Mean (SD) [range] | 5.7 (2.5) [2.8 – 11.7] | 6.6 (2.3) [2.8 – 10.5] | 0.112<sup>b</sup> |
| **IQ**<br>Mean (SD) [range] | 73.0 (24.1) [43 - 121] | 75.8 (22.1) [40 -115] | 0.639<sup>b</sup> |
| **SES**<br>Mean (SD) [range] | 54.0 (19.1) [30 - 90] | 53.1 (20.7) [10 - 90] | 0.864<sup>b</sup> |
| **TIV (cm<sup>3</sup>)**<br>Mean (SD) | 1438.2 (15.1) | 1426.3 (15.7) | 0.748<sup>b</sup> |
| **ADOS - Total**<br>Mean (SD) [range] | 6.16 (1.77) [4 - 10] | - | - |
| **ADOS - SA**<br>Mean (SD) [range] | 6.23 (1.85) [3 - 10] | - | - |
| **ADOS - RRB**<br>Mean (SD) [range] | 6.87 (1.57) [5 - 10] | - | - |

### 3.2 Social-communication and motor measures

With respect to the social-communication domain, autistic participants had higher scores on social cognition, social communication and autistic mannerisms subscales of SRS compared to non-autistic peers (Table 2). Considering motor skills, the autistic group scored lower in the motor control and coordination subscales of DCDQ (Table 3).

**Table 2.** Social-communication measures, subscales of the SRS, in autistic and non-autistic participants.

| **SRS subscales** | **Autistic**<br>n=30<br>mean (SD) | **Non-autistic**<br>n=29<br>mean (SD) | **p-value** |
| --- | --- | --- | --- |
| Social awareness | 10.1 (3.5) | 8.3 (3.6) | 0.051 |
| Social cognition | 16.0 (7.8) | 11.1 (5.2) | **0.006** |
| Social communication | 27.0 (11.1) | 18.1 (9.3) | **0.002** |
| Social motivation | 12.8 (5.1) | 10.0 (6.6) | 0.070 |
| Autistic mannerisms | 14.2 (7.1) | 8.7 (5.4) | **0.001** |

**Table 3.** Motor measures, subscales of the DCDQ, in autistic and non-autistic participants.

| **DCDQ subscales** | **Autistic**<br>n=30<br>mean (SD) | **Non-autistic**<br>n=30<br>mean (SD) | **p-value** |
| --- | --- | --- | --- |
| Motor control | 14.7 (4.6) | 18.4 (6.2) | **0.013** |
| Fine motor | 8.2 (4.2) | 9.7 (5.0) | 0.214 |
| Coordination | 11.8 (3.8) | 14.6 (5.2) | **0.023** |

### 3.3 Neuroanatomical measures

No statistically significant difference was found in total CC volume between autistic and non-autistic participants (Table 4). Considering the volumes of the 5 CC subregions, the one-way ANOVA revealed significantly smaller volumes in the central subregion for the autistic group (F(1,68)=11.0, p=.001, η^2^= 0.139; Cohen’s d= −0.792, 95% CI [-1.29,-0.296]). No significant differences were found for the other subregions of the CC (Figure 1). The between-group difference in the volume of the central CC remained significant after controlling for TIV, or for TIV and age (Table 4).

**Table 4.** Volume of the CC (in mm3) and its 5 subregions according to Freesurfer parcellation in autistic and non-autistic participants. TIV = total intracranial volume.

|  | Autistic<br>n=35<br>mean (SD) | Non-autistic<br>n=35<br>mean (SD) | Group difference (p-value) |  |  |
| --- | --- | --- | --- | --- | --- |
|  |  |  | Unadjusted | TIV as covariate | TIV and Age as covariates |
| **Total CC** | 3018.8<br>(616.2) | 3138.5 (601.2) | 0.414 | 0.228 | 0.299 |
| **Posterior CC** | 758.3 (149.7) | 733.4<br>(162.0) | 0.507 | 0.564 | 0.385 |
| **Mid Posterior CC** | 430.4 (123.3) | 431.8<br>(117.2) | 0.961 | 0.818 | 0.839 |
| **Central CC** | 486.4 (132.1) | 596.7<br>(146.1) | **0.001** | **<0.001** | **0.001** |
| **Mid Anterior CC** | 506.8 (151.4) | 562.9<br>(163.4) | 0.141 | 0.079 | 0.076 |
| **Anterior CC** | 837.0 (208.6) | 813.7<br>(167.2) | 0.608 | 0.679 | 0.833 |

**Figure 1.**
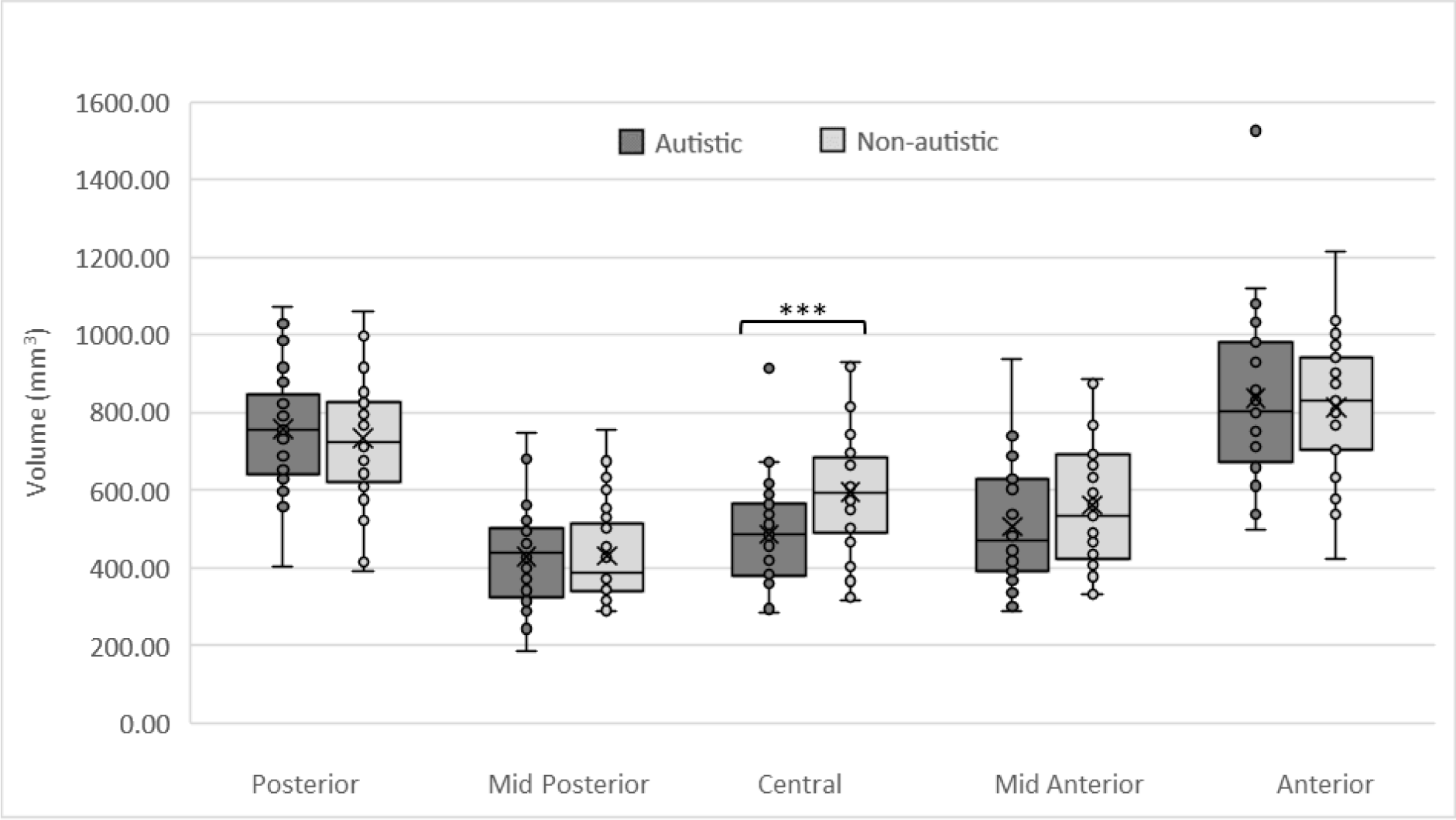
Volumes of the 5 CC subregions in autistic and non-autistic participants. Significant differences in volume between groups are seen in the central subregion (***, p=.001).

### 3.4 Correlations between CC and social-communication/motor measures

Across participants, central CC volume was significantly associated with two DCDQ subscales —motor control, and fine motor, namely— (Pearson’s r =0.365, p=0.004, and Pearson’s r =0.406; p=0.001, respectively; Figure 2, panels A and B). Pearson’s correlation analysis also revealed a significant negative association between central CC volume and the SRS subscale autistic mannerisms (r= −0.280; p=0.032; Figure 2, panel C). When controlling for TIV, only the correlations with the two DCDQ subscales remained significant (DCDQ motor control: r=0.292; p=0.025; DCDQ fine motor: r= 0.296; p=0.023). After controlling for TIV and age, only the correlations between central CC volume and DCDQ motor control (r= 0.270; p=0.040) and SRS autistic mannerisms (r= −0.262; p=0.049) continued to be significant (DCDQ fine motor showed a trend to significance: r= 0.251; p=0.057).

**Figure 2.**
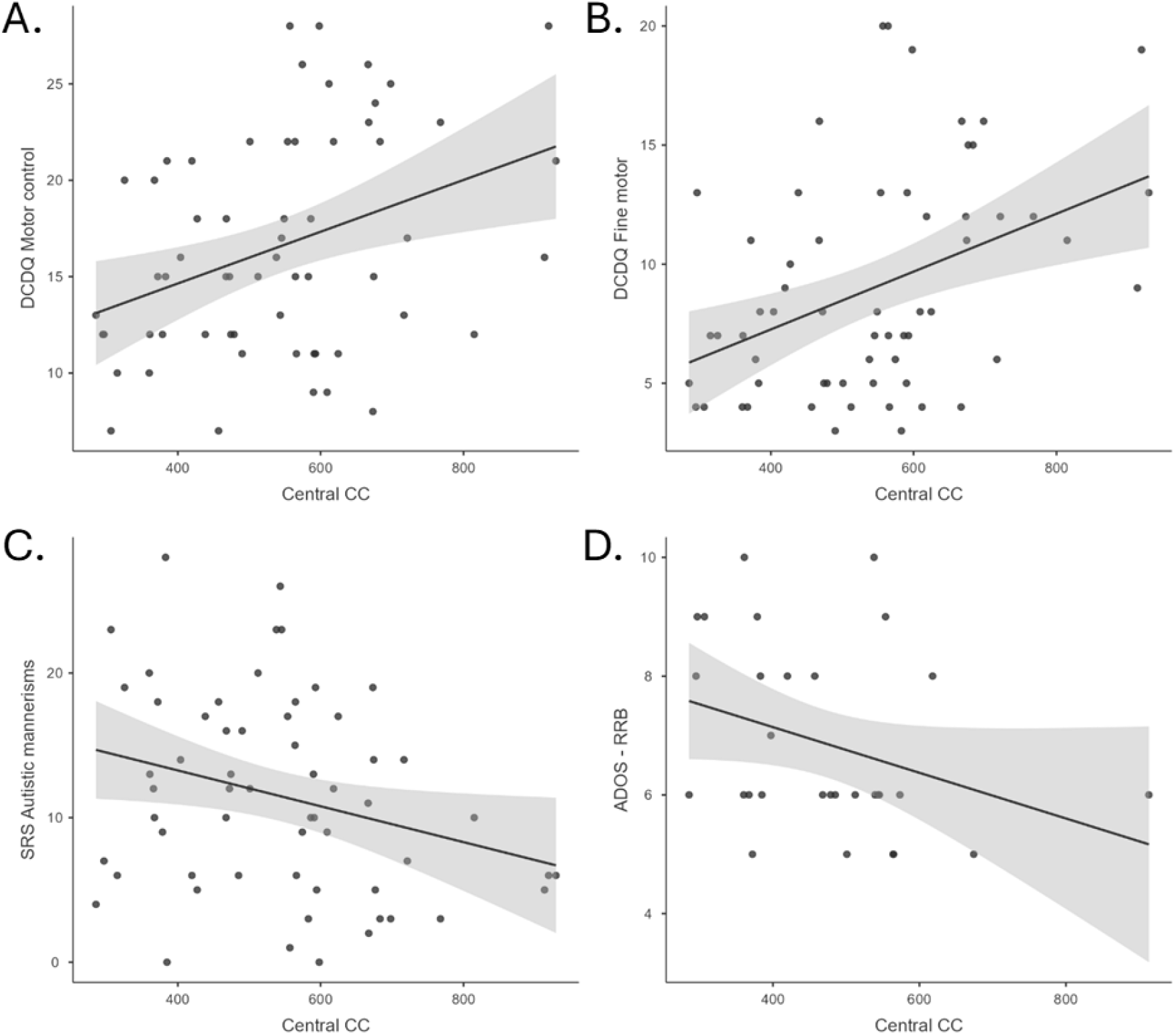
Significant correlations between central CC volume and social-communication/motor measures: DCDQ subscales motor control (panel A) and fine motor (panel B), SRS subscale autistic mannerisms (panel C), and ADOS subscale restricted repetitive behaviours (RRB; panel D).

Finally, Spearman’s correlations of central CC volume with the ADOS subscales showed a significant negative correlation (r= −0.370; p=0.044) with ADOS-RRB (figure 2, panel D).

## 4. DISCUSSION

The present study sought to investigate volumetric differences in the CC between autistic children and a comparison group of non-autistic peers in the same IQ range. Furthermore, we aimed to explore the possible relationship between CC volume and core and non-core features of autism. The main finding is that autistic children show a reduced volume of the central segment of the CC in the context of a comparable CC total volume. This reduction appears to be correlated with restricted and repetitive behaviours symptoms and to parental ratings of motor skills.

A widespread decrease in callosal size is considered to be a well-replicated finding in autism (Frazier & Hardan, 2009; Valenti et al., 2020), although not always supported by large-sample investigations (Lefebvre, Beggiato, Bourgeron, & Toro, 2015). The finding of statistically comparable CC’s total volume between autistic and non-autistic participants in the present study is indeed in line with several previous reports (Cheng et al., 2010; Giuliano et al., 2018; Haar, Berman, Behrmann, & Dinstein, 2016; Loomba, Beckerson, Ammons, Maximo, & Kana, 2021; Mengotti et al., 2011). Nevertheless, a specific subregion of the CC —the central CC— was found to be significantly reduced in our sample of autistic participants. This finding is in line with other studies which reported a conserved total CC in spite of a significantly reduced central CC in autistic subjects from the large ABIDE database (Haar, Berman, Behrmann, & Dinstein, 2016; Loomba, Beckerson, Ammons, Maximo, & Kana, 2021). The reduction in CC volume is coherent with the hypothesis of underconnectivity, at least in the networks including this section of the CC (Zito et al., 2014; Just, Cherkassky, Keller, Kana, & Minshew, 2007).

The central segment of CC has been previously associated with motor functions. Diffusion imaging findings about the topographical organisation of the CC indicated that the fibres connecting contralateral cortical areas involved in motor planning and execution (e.g., primary motor, supplementary motor, and premotor cortices) pass through the central segment of the CC (Hofer & Frahm, 2006; Russo et al., 2022; Zarei et al., 2006). Our exploratory correlational analyses provide further support to this evidence. Indeed, across participants, the volume of the central CC was associated with motor abilities as assessed by the DCDQ questionnaire, specifically in the domain of motor control. Interestingly, this domain includes items referred to ball skills (such as throwing, catching or hitting a ball), which are among the most documented motor features in autism (De Francesco et al., 2023; Lidstone & Mostofsky, 2021; Whyatt & Craig, 2012). The correlation in our data seems therefore to suggest that an increase of volume of the white-matter tracts likely connecting motor-relevant structures corresponds to better motor performance as judged by parents. Coherently, a larger CC size has been previously related with better motor development in healthy at-term infants (Chang, Hung, Yang, Ho, & Chiu, 2015), as well as with higher motor proficiency as assessed by the Movement Assessment Battery for Children in school-aged children and adolescents born preterm (Lubián-Gutiérrez et al., 2024; Rademaker et al., 2004; Van Kooij et al., 2008). However, this result differed from previous reports in autism that did not find any significant association between CC volume and direct motor assessments (Freitag et al., 2009; Hanaie et al., 2014). This discrepancy could be due to the fact that, in the current work, we recruited autistic participants with lower IQ scores and younger ages than the ones in Freitag et al.’ and Hanaie et al.’ studies. This study also differs in other methodological aspects from previous ones, given that Freitag and colleagues assessed the global size of the CC, whereas Hanaie and colleagues evaluated the CC body tract volume as reconstructed by diffusion tensor imaging (DTI) (Freitag et al., 2009; Hanaie et al., 2014). To note, the present study included a larger group of participants than previous works.

Furthermore, when the measures of core autistic traits/symptoms were considered, we found a significant relationship between the reduction of the volume of the central CC and higher levels of mannerisms across participants, and restricted repetitive behaviours in the autism group, respectively. The association with measures of stereotyped behaviours has been previously reported (Hardan et al., 2009), as well as a predictive association of early CC size with later presentation of repetitive behaviours (Wolff et al., 2015). Thus, although exploratory, the present findings provide further empirical evidence that the reduced connectivity through CC might be relevant for the basic mechanisms underpinning also the core features of the autistic condition.

Interestingly, we did not find any associations between the central CC size and social subscales of the SRS across participants. This appears to be in contrast with a previous report (Loomba, Beckerson, Ammons, Maximo, & Kana, 2021), which found a significant negative correlation between central CC volume and total SRS scores. Nevertheless, the lack of significant associations with parental ratings of social functioning, as well as with the social affect subscale of the ADOS, could provide support to the idea that motor atypicalities (but also restricted and repetitive behaviours) represent a dimension that is orthogonal to the core social features of autism (Mandelli et al., preprint). According to this hypothesis, future extensions of the present work should consider using motor features to stratify the autistic condition in different subgroups and then investigate potential neuroanatomical differences.

This work has several limitations that need to be addressed. First, although similar to previous lab specific works, our sample size is small compared with the neuroanatomical database used in more recent studies (i.e., the ABIDE). Second, the present results may not be generalizable to a larger autistic population given that our sample was clinically referred and included participants who achieved low IQ scores. Conversely, this could also be considered a strength of our work, as only few imaging studies comprised autistic participants with lower scores at standardized measures of IQ. Likewise, 15 out of 35 children in our comparison group also presented an idiopathic intellectual disability. Although intellectual disability can be overall associated with alterations in brain morphology (Ma et al., 2021), matching participants on IQ represents a viable option to reduce the influence of this factor (Jack & Pelphrey, 2017). Moreover, MRI scans were carefully inspected by an expert child neuroradiologist (FA) to exclude any major brain abnormality across all sample. Even so, it is important to acknowledge that the absence of a specific cause for the intellectual disability in some participants of the comparison group did not guarantee that these children form the lower tail of the typical IQ distribution within the general population (see Jarrold & Brock, 2004). A further limitation of this work is the lack of standardised assessments of motor skills besides parent-report questionnaires. Even though these latter could be relevant to address the adaptive implications of motor skills on daily life, questionnaires still represent a partial point of view, which could be enriched by direct motor assessments and/or kinematic recordings of the movements. Lastly, the segmentation of the CC in different subregions used here was based on its length rather than on the specific identification of the existent tracts functionally connecting specified cortical areas, such as done by using tractography techniques (Huang et al 2005; Wahl et al. 2007). Nevertheless, multiple studies demonstrated that, in a typical anatomy, the fibres connecting the cortical areas involved in motor planning and execution across the two hemispheres are located in the central section of the CC (Hofer & Frahm, 2006; Wahl et al. 2007; Park et al 2008; Fabri et al 2014).

## 5. CONCLUSIONS

In summary, the present study provides evidence that a group of drug-naïve autistic children aged 2.5-12 years showed a decreased central CC volume in comparison with a group of closely IQ-matched, non-autistic peers. Furthermore, smaller central CC volumes were associated with restricted repetitive behaviours in autistic participants, and to the degree of mannerisms and motor control across participants. Overall, these findings contribute to expanding the current knowledge about the neural mechanisms underlying both core and non-core features of autism, possibly supporting the underconnectivity hypothesis of the condition.

## Acknowledgments

The authors acknowledge the work of prof Renato Borgatti and all the clinical team at Neuropsychiatry and Neurorehabilitation Unit, Scientific Institute, IRCCS Eugenio Medea, (Bosisio Parini, Lecco) in the diagnostic evaluation of participants referred for suspected developmental delays. The authors are also especially grateful to all the families of the children who took part in this study.

## Conflict of interest

The authors declared no potential conflicts of interest with respect to the research, authorship, and/or publication of this article.

## FUNDING INFORMATION

This work was supported by grants from the Italian Ministry of Health to AC (Ricerca Finalizzata GR-2011-02348929; Ricerca Corrente 2024) and from the Italian Ministry of University and Research to EM (grant n. 2022RXM3H7).

## Data availability statement

The data that support the findings of this study are available on request from the corresponding author. The data are not publicly available due to privacy or ethical restrictions.

## Notes

### Competing Interest Statement

The authors have declared no competing interest.

### Author Declarations

The Ethics Committee of our Institute Comitato Etico IRCCS E. Medea - Sezione Scientifica Associazione La Nostra Famiglia gave ethical approval for this work (Prot. N.33/18 - CE).

